# Diagnostic criteria for patellofemoral pain syndrome: a scoping review protocol

**DOI:** 10.1101/2023.03.25.23287736

**Authors:** Alberto Casalini, Davide Morganti, Angela M.R Salvemini

**Affiliations:** Usl Toscana Sud Est, C.A.R.F Tamburino, Siena, Italy; Department of Medicine, Surgery and Neuroscience, University of Siena; Muscoloskeletal and rheumatological physiotherapy, Department of Human neurosciences, University of Roma “Sapienza”

## Abstract

**Background:** Knee pain is a frequent cause of mobility limitation and worsening quality of life. There are several atraumatic musculoskeletal disorders clinically presenting with knee pain and among them, there is patellofemoral pain syndrome (PFPS). According to the biopsychosocial approach, it is important to consider other factors and the whole person besides the anatomical structure. A specific diagnosis is important when a macroscopic damage is present and can be investigated with accurate test and diagnostic imaging. This scoping review aims to provide an overview of clinical tests and imaging commonly used in clinical practice to identify the aforementioned atraumatic knee pathology.

**Inclusion criteria:** Studies considering patients with patellofemoral pain syndrome (PFPS) diagnosis and, in particular, studies concerning the diagnosis of this disorder in a rehabilitation and medical setting, will be included.

**Methods:** This review will follow the Joanna Briggs Institute (JBI) methodology. Medline, Scopus, Pedro will be searched. In addition, other records will be identified searching grey literature. Limits regarding year of publication will not applied. Secondary research studies will be excluded and only English studies will be included. Two reviewers will independently screen all studies for inclusion and disagreements will be resolved by a third author. A data collection table will be developed by the researchers and a narrative and graphical synthesis of the results will be presented.

**Conclusion:** This scoping review will be the first presenting an overview of criteria used for the diagnosis of the PFPS. The results will help clinicians to consider the role of clinical tests and imaging in the framing of patients with patellofemoral pain syndrome (PFPS).

## Introduction

Knee pain is a frequent cause of mobility limitation and worsening quality of life in 25% of adults (1). Musculoskeletal pain disorders account for about 20-30% of first visits to the emergency department; of these, shoulder, back, and knee issues are the most prevalent.

Anterior knee pain or dysfunction is a common presentation and reason for specialist visits (2). There are several atraumatic musculoskeletal disorders clinically presenting with knee pain, and among them, there are patellofemoral pain syndrome (PFPS), iliotibial band syndrome (ITBS), and Hoffa’s disease. Patellofemoral pain syndrome prevalence in the general population is 22.7 %, with a prevalence in women of 29.2 %, in men of 15.5 %(3) and in the adolescent population of 28.9 % (4).

Patellofemoral syndrome incidence rate in female amateur runners is 1080.5/1000 person-years(5). Biopsychosocial approach emphasizes how important is to consider the whole person rather than the pathology itself and the importance of a specific diagnosis only in cases where the anatomic structure has macroscopic damage that can be investigated with accurate tests and diagnostic imaging. Therefore, the need to map the literature about diagnostic criteria, including clinical tests, subjective history and imaging methods, commonly used to investigate anatomic structures correlated with patellofemoral pain syndrome.

To the authors’ knowledge, there are four systematic reviews which aim to investigate the diagnostic accuracy of patellofemoral pain syndrome clinical tests (6–9)

This scoping review aims to provide an overview of diagnostic criteria commonly used in clinical practice to identify patients with PFPS.

Currently, in clinical practice, clinicians use imaging and clinical tests to complete the anamnesis and physical examination and determine a diagnostic hypothesis, often based solely on the structure, which is identified as the main cause of the patient’s symptoms. This is what also occurs in atraumatic conditions such as PFPS. In order to provide an overview of clinical tests and imaging methods used for the diagnosis of the above pathology, a scoping review will be conducted, which is the most appropriate study design to map the evidence in the literature and identify any gaps in research. (10) This review is intended to be a starting point for future studies investigating the role of tests and imaging in the framing of patients with patellofemoral pain syndrome.

## Methods

This scoping review will follow the Joanna Briggs Institute (JBI) methodology. (10) for the conduct and reporting of scoping reviews. The Preferred Reporting Items for Systematic reviews and Meta-Analyses extension for Scoping Reviews (PRISMA-ScR) Checklist for reporting will be used (11).

### Inclusion criteria

#### Participants

The review will consider all studies including patients presenting with patellofemoral pain syndrome. This is a patient population clinically presenting with knee pain unrelated to traumatic causes. Studies targeting patients with associated traumatic nature injuries, patients with rheumatic diseases, patients undergoing previous surgery, and patients with severe arthrosis grade III and IV of the Kellgren Lawrence scale and positive for the *ACR Clinical Classification Criteria for Osteoarthritis of the knee diagnostic criteria*(12) will be excluded. In addition, cadaver studies will be excluded.

#### Concept

The present review will consider all studies that use any criteria for the diagnosis of patellofemoral pain syndrome. In particular, all studies describing clinical tests, subjective history and imaging methods useful in identifying the aforementioned disorder will be included.

#### Context

This scoping review aims to investigate the literature regarding the clinical /radiographic diagnosis of the patellofemoral pain syndrome in a rehabilitation and medical setting. All studies meeting the inclusion criteria will be included in order to provide a broad overview of the available evidence.

### Types of sources

We will consider primary research studies from 1952 to the present, like randomized controlled trial, non-randomized controlled trial, prospective and retrospective cohort studies, case control studies, analytical cross sectional studies, case series, case report and descriptive cross-sectional studies. In addition, also grey literature will be searched. Studies not meeting inclusion criteria and study designs like systematic reviews, narrative reviews, scoping reviews, opinion papers and letters will be excluded.

### Search strategy

An initial search on Medline was conducted to identify keywords and MeSH terms used to describe the topic. Identified terms was used to develop a search strategy for Medline (see **Table 1**). Search strategy and identified keywords will be adapted for the following database: Scopus, PEDro. In addition, grey literature will be searched (e.g Google scholar, OpenGrey).

**Table 1.**
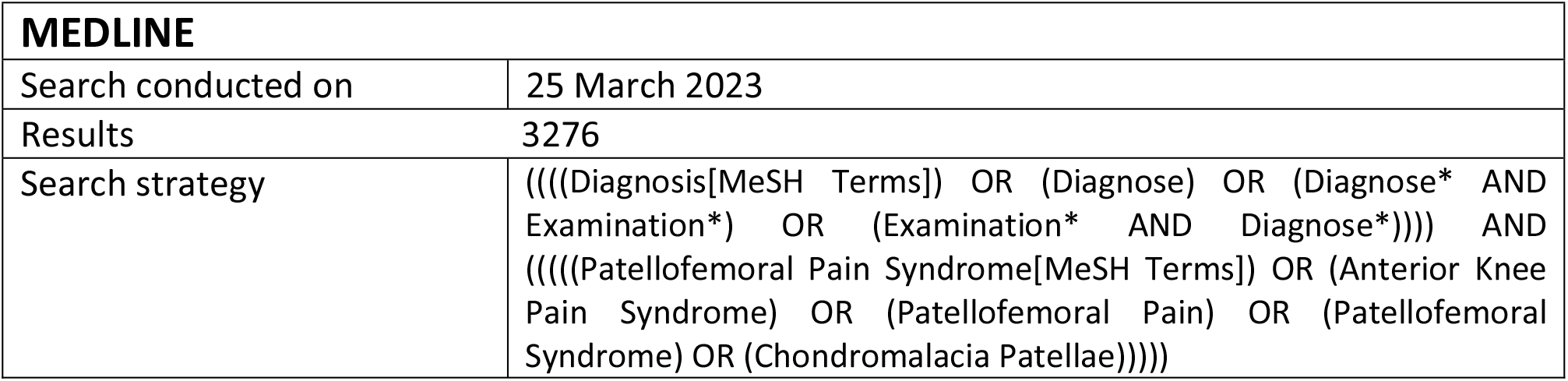
Search strategy for Medline

**Table 2.**
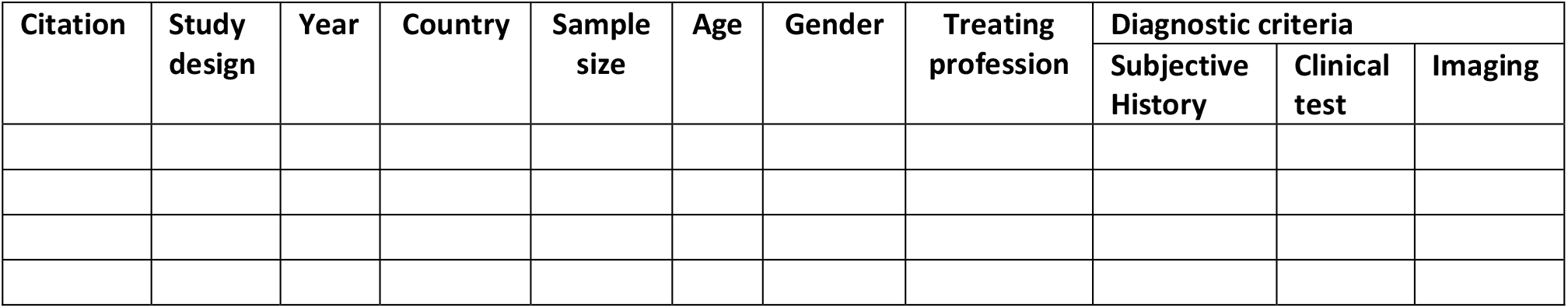
Data extraction

### Study selection

Screening will be performed by two independent authors via Rayyan QCRI online software (13).

The first screening will be performed by title and abstract and then by reading the full text. Conflicts will be resolved by a third author. The study selection process will be summarized through the latest published version of Preferred Reporting Items for Systematic Reviews and Meta-analysis (PRISMA) flow diagram. (14) Excluded articles will be collected in the scoping review report and the reason about their exclusion will be reported.

### Data extraction

A table (Table2) will be developed by the reviewers that will descriptively collect key information and data from the considered studies. Two authors will be involved in data extraction. The extraction will occur independently with a final cross-checking. Disagreements will be resolved by a third author. Following data will be extracted and presented:

- author
- year of publishing
- country origin of study
- sample size
- age
- gender
- treating profession
- criteria used for the diagnosis of PFPS such as subjective history, clinical tests, imaging
- pain site
- symptom duration
- study design

Any further modifications/integrations will be detailed in the full scoping review.

### Data presentation

The results of the scoping review will be presented in a descriptive narrative synthesis that relates to the aims and objectives of the review. Quantitative data will be presented descriptively using percentages and graphical representations will be used. In particular, diagnostic criteria will be divided in three categories: clinical tests, subjective history and imaging. Every category will be presented with a histogram representing with percentages how many studies investigate a specific criteria. Results from the histograms will be collected in a table showing the most common diagnostic criteria used for the diagnosis of patellofemoral pain syndrome.

A discussion will be provided regarding the results of the scoping review. Final conclusions will be drawn from the mapped evidence.

## Data Availability

All data produced in the present study are available upon reasonable request to the authors

## Acknowledgements

Not applicable.

